# Quantifying the role of importation on sustained malaria transmission in a low-to-moderate burden region of Southwest Uganda

**DOI:** 10.1101/2025.05.14.25327608

**Authors:** Adrienne Epstein, Okiria Aramanzan, Isaiah Nabende, Tonny Max Kayondo, Michael Obbo, Robert Tumwesigye, Innocent Wiringilimaana, Monica Mbabazi, Brian A Kagurusi, Stephen Tukwasibwe, Isaac Ssewanyana, Isobel Routeledge, Jessica Briggs, Amy Wesolowski, Bryan Greenhouse, Grant Dorsey, Emmanuel Arinaitwe, Isabel Rodriguez-Barraquer

**Affiliations:** Department of Medicine, University of California San Francisco, San Francisco, California, USA; Infectious Diseases Research Collaboration, Kampala, Uganda; Department of Epidemiology, Johns Hopkins School of Public Health, Baltimore, Maryland, USA; Chan Zuckerberg Biohub, San Francisco, CA, USA

## Abstract

**Background:** Parasite importation remains a critical challenge to malaria elimination efforts. The extent to which human travel contributes to sustained transmission in low-to-moderate burden areas of highly endemic African countries is poorly understood.

**Methods:** We conducted a 14-month longitudinal cohort study in Kamwezi subcounty in Southwest Uganda, an area targeted for malaria elimination where sustained transmission poses challenges. A total of 1,918 individuals from 400 households were enrolled and followed bi-monthly. Travel histories, malaria episodes, and household characteristics were collected through structured surveys. Incident malaria was captured via passive case detection at the local health facility and self-report. Multilevel logistic regression models estimated the association between overnight travel and incident malaria, adjusting for demographic and household factors. Population attributable fractions (PAF) quantified the contribution of travel to malaria burden, stratified by transmission intensity, season, and village of residence.

**Results:** Over the study period, 244 malaria episodes were recorded, with incidence highest in villages with greater baseline transmission. Nearly one-third of participants reported at least one overnight trip. We found that associations between travel and malaria varied spatially and temporally, with positive associations in villages of lower transmission (odds ratio=5.65, 95% CI: 2.06-15.45) and during periods of low overall malaria transmission. We found these associations to be strongest for short-distance trips to nearby areas of higher incidence. PAF analyses suggested that travel accounted directly for 17% of malaria cases in low-transmission villages overall, rising to 44.6% during periods of low transmission.

**Conclusion:** Overnight travel contributed meaningfully to malaria burden, especially in villages with low local burden and during periods of seasonally low transmission. These effects were evident even at small geographic and temporal scales. These findings underscore the importance of malaria surveillance and control strategies that account for both local transmission and travel-related importation, even outside of elimination contexts.

## Introduction

Significant progress has been made in reducing malaria burden in sub-Saharan Africa over the past two decades, with malaria incidence declining by 36% and mortality by 29% between 2000 and 2023.^1^ Despite these achievements, malaria remains a leading cause of morbidity and mortality in the region, with an estimated 246 million cases and 569,000 deaths reported in 2023 ^1^. As malaria burden decreases and becomes increasingly spatially heterogeneous – driven by the scale-up of vector control interventions and improved case management – importation of malaria parasites has emerged as a critical challenge to control and elimination efforts. Travel to and from malaria-endemic areas may compromise local control measures by introducing parasites which may sustain transmission in affected areas.^2-6^ Though often discussed, the rate of parasite importation remains largely speculative due to the lack of comprehensive studies conducted in natural settings across sub-Saharan Africa.

Uganda is illustrative of a country that has substantial heterogeneity in malaria transmission, due to geographic targeting of control measures and natural differences in environmental characteristics across the country. While the national malaria burden remains high, the Ministry of Health has set subnational elimination targets based on areas’ receptivity and transmission characteristics.^7^ For example, the Kigezi sub-region in the southwest demonstrated the lowest parasite prevalence nationally in the 2018-2019 Malaria Indicator Survey (<1% among children aged 0–59 months)^8^ and is targeted for malaria elimination in the Uganda National Malaria Strategic Plan 2025.^9^ However, even within Kigezi, malaria burden is unevenly distributed, with hotspots of high malaria incidence posing challenges to elimination efforts. Given these factors, there is limited understanding of the extent to which parasite importation contributes to sustained transmission in this region.

This study aimed to quantify the impact of importation on malaria burden over a 14-month period in a community within Kamwezi subcounty, Kigezi sub-region, Uganda. Using data from an ongoing longitudinal cohort study of 400 households combined with passive case detection at a public health facility, we characterized the association between overnight travel and incident malaria and estimate the proportion of malaria cases that are attributable to travel, considering key factors such as malaria burden at the travel destination and local malaria burden at the time of travel. Finally, we aimed to estimate the attributable risk of overnight travel on malaria within this population and assess potential variation in attributable risk spatially and temporally.

## Methods

### Setting

This cohort study was conducted in a 35 square-kilometer area in Kamwezi subcounty, Rukiga District, Uganda, along the border with Rwanda (Figure 1). The area has low-to-moderate malaria transmission with substantial spatial and temporal variation. Since 2013, Uganda’s Ministry of Health has implemented national long-lasting insecticidal net (LLIN) distributions every 3-4 years; the most recent in Kamwezi occurred in November 2023 using chlorfenapyr and alpha-cypermethrin LLINs. Indoor residual spraying (IRS) has never been implemented in this area. These analyses use 14 months of follow-up data (seven household visit rounds from July 2023 to September 2024).

**Figure 1.**
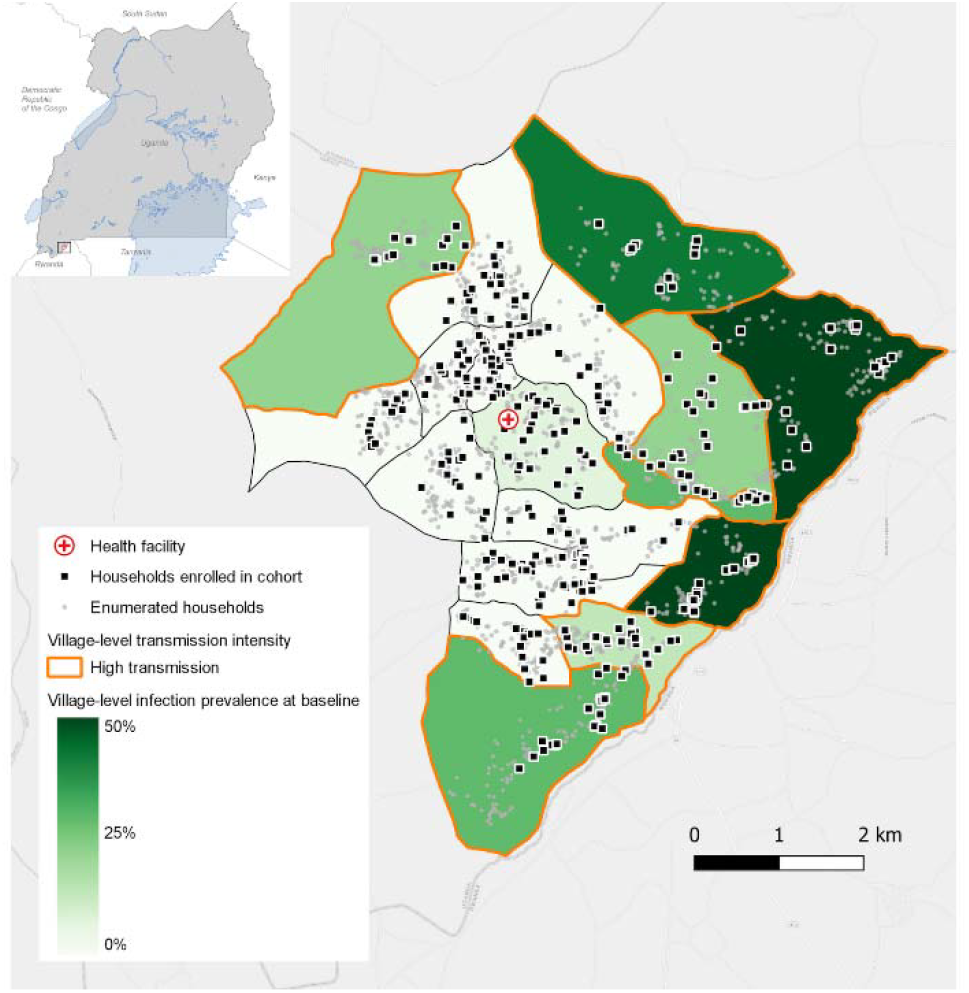
Map of households enrolled in cohort and village-level *Plasmodium falciparum* infection prevalence by qPCR.

### Screening and enrollment of cohort households

In January 2023, all households within a geographically defined study area were enumerated and mapped (n=2,904) using handheld global positioning systems to create a sampling frame for household recruitment (see Supplemental Figure 1 for cohort profile diagram). Households were defined as single permanent or semi-permanent dwelling structures serving as the primary residence for individuals or groups who cook and eat together. After enumeration, village-level household enrollment targets were set proportional to the number of households in each village. Stratified random sampling was then performed based on these targets to select households for screening and enrollment. Households were enrolled if they met the following criteria: 1) at least one adult aged 18 years or older was present; 2) at least 80% of household members were willing to participate; 3) an adult resident agreed to provide informed consent for the household survey; and 4) there were no plans for the household to relocate from the study area within the next two years.

### Screening, enrollment, and follow-up of cohort participants

All full-time residents of enrolled households (defined as individuals intending to have their primary sleeping place at that location for the next six months) were screened and enrolled in the longitudinal cohort study beginning in July 2023 if they met the following criteria: 1) they were permanent residents of the household, and 2) they provided written informed consent (from a parent or guardian for children aged 6 months to 17 years). The cohort was dynamic, allowing for the screening and enrollment of any new permanent residents joining a household over the course of the study.

Enrolled households were visited every two months for follow-up. During each visit, the head of household (or a designate) completed a questionnaire covering household characteristics, malaria control interventions, and details about visitors in the preceding two months, including approximate dates of visits, reasons for visits, and the visitors’ origins. All household participants also completed surveys about malaria illness and treatment over the past two months, visits to health facilities, and usage of LLINs. In addition, participants provided detailed information about travel, including destinations, the number of nights spent away, and malaria control interventions used during travel. A small blood sample (<1 mL via finger prick, collected into microtainers and as dried blood spots) was also collected from participants at all visits. For this analysis we use data from 14 months (7 visits) through September 2024, unless a participant was prematurely withdrawn due to 1) permanent relocation outside the study area, or 2) withdrawal of informed consent.

### Passive health facility-based surveillance

In parallel to the cohort, passive surveillance at the local health facility (shown the map in Figure 1) was conducted by enrolling patients with laboratory-confirmed *Plasmodium falciparum* (either through rapid diagnostic test or microscopy). If participants were members in the longitudinal cohort study, their subject identifier was recorded and a travel survey like that delivered in the cohort study was administered.

### Laboratory procedures

DNA was extracted from all samples from the household study using the Tween-Chelex method, as previously described.^10^ Quantitative PCR (qPCR) was performed on all samples from the household study to determine the presence/absence and parasitemia in parasites per microliter of *Plasmodium falciparum* parasites. Extracted products were tested for the presence and quantity of *P. falciparum* DNA via a sensitive *var* gene acidic terminal sequence qPCR assay targeting the multicopy conserved *var* gene acidic terminal sequence.^11^ A standard curve consisting of DNA extracted from dried blood spots with cultured, ring stage parasites diluted in whole blood to parasite densities ranging from 1 to 10,000 parasites/μl was included in duplicate on each plate.

### Variable definitions

#### Importation exposure variables

While importation of *P. falciparum* can occur through various types of travel, for this study we focused on overnight travel given the nocturnal biting behavior of the most common *Anopheles* vectors. Recent overnight travel was defined as spending at least one night in a village outside the village of residence within the prior two months. We evaluated several overnight travel-related exposure variables and their association with malaria illness: 1) a binary variable representing whether the respondent reported any overnight travel; 2) a binary variable representing whether the respondent reported any overnight travel to an area with higher district- or, when available, parish-level incidence (defined using the Malaria Atlas Project’s 2020 incidence estimates^12^) than the village of residence; and 3) travel duration in weeks (categorical; no travel, less than 1 week, greater than 1 week). For those participants who reported going to areas of higher incidence we also evaluated 4) travel destination at the district level and 5) travel distance (categorical; no travel to areas of higher incidence, less than 25 kilometers, 25-50 kilometers, and greater than 50 kilometers). For cases detected at the health facility, we used travel histories captured during the health facility visit; for all other individuals, we used travel histories conducted at bi-monthly household surveys.

#### Outcome

The outcome for all analyses was individual-level incident malaria. Individuals were classified as having an incident episode of malaria if they were diagnosed with malaria at the study health facility in the two months preceding the survey. In addition, during routine visits individuals answered the yes/no question “Have you had malaria in the last two months?” and those individuals who answered yes were classified as having had an incident episode even if they were not captured at the health facility. We conducted sensitivity analyses considering only confirmed malaria cases detected at the health facility as outcomes; these analyses excluded baseline data because we were not able to confirm cases prior to enrollment in the study.

#### Covariates

We adjusted models for covariates that may influence both the likelihood of travel and malaria risk: 1) participant gender; 2) participant age (categorical; <5, 5-15, and >15); 3) a binary variable representing improved walls in the participant’s household (walls made of burnt bricks, cement blocks, stone, timber, or iron sheets versus thatch, mud, or unburnt bricks); 4) a binary variable representing improved floors in the participant’s household (floors made of parquet or polished wood, tiles, bricks, cement/concrete, or stones versus earth, sand, or dung); 5) a numeric variable representing the number of household residents; and 6) an indicator variable representing calendar time to account for seasonal changes in travel patterns and malaria risk.

### Data analysis

We used multilevel logistic regression models to estimate odd ratios (ORs) and 95% confidence intervals (CIs) for the relationship between travel-related variables and incident malaria. Our primary analysis included models for individuals living in all villages and models stratified by transmission intensity (high vs. low transmission villages). Transmission intensity was defined using village-level prevalence of parasitemia at baseline measured via qPCR. High transmission villages were defined as those with a prevalence of at least 10% (the median prevalence across all villages). Additionally, we assessed whether the associations between travel variables and incident malaria changed over time and further examined whether these associations varied by both time and village-level transmission intensity. For models including all villages, we incorporated both village- and individual-level random effects. For models stratified by transmission intensity, we used individual-level random effects only. As a secondary analysis, we varied the threshold used to classify the transmission intensity (high vs. low) using thresholds ranging from 5% to 50%. For this analysis we allowed the classification of each village to vary over time based on village-level prevalence measured throughout follow-up visits every two months. This analysis estimated the association between travel-related variables and incident malaria across villages above and below each threshold.

We estimated population attributable fractions (PAF) and their 95% CI using the formula 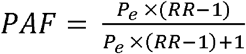 where *P*_*e*_ is the prevalence of the exposure in the population, and RR is the relative risk of the travel-related exposure and incident malaria. Here, odds ratios represented the ratio of risk for these associations. We estimated PAFs overall, by transmission intensity, season, village, and demographics. The PAFs resulting from these analyses represent the impact of direct importation on malaria burden, not including any secondary infections that may result from transmission downstream of an imported case.

## Results

### Study population characteristics

A total of 1,918 individuals residing in 400 households were enrolled in the cohort and included in this analysis (Table 1). More than half of participants were female, and half were older than 15 years of age. The most common occupation among those older than 18 years of age was farmer. Approximately one-quarter of participants had no formal education, while more than half completed primary-level schooling, and the rest completed secondary school or beyond. Seven hundred eighty-four participants were from high transmission villages and 1,134 from low transmission villages, and demographics were similar between these two types of settings.

**Table 1.** Characteristics of cohort participants at baseline (or first time surveyed), overall and stratified by village-level transmission intensity.

|  | All villages (n=1,918) | High transmission villages* (n=784) | Low transmission villages (n=1,134) |
| --- | --- | --- | --- |
| Female, % (n) | 57.2 (1,097) | 58.9 (462) | 56.0 (635) |
| Age, % (n) |  |  |  |
| <5 | 15.0 (288) | 16.3 (128) | 14.1 (160) |
| 5-15 | 35.1 (674) | 36.1 (283) | 34.5 (391) |
| >15 | 49.8 (956) | 47.6 (373) | 51.4 (583) |
| Occupation (among those greater than 18 years), % (n) |  |  |  |
| Commerce | 6.0 (49) | 6.8 (21) | 5.6 (28) |
| Farmer | 76.4 (620) | 83.2 (257) | 72.2 (363) |
| Student | 5.3 (43) | 4.5 (14) | 5.8 (29) |
| Childcare/homemaker | 1.6 (13) | 0.3 (1) | 2.4 (12) |
| Other | 10.5 (85) | 5.2 (16) | 13.7 (69) |
| Education level |  |  |  |
| None | 24.4 (468) | 26.7 (209) | 22.8 (259) |
| Primary | 54.8 (1,052) | 54.3 (426) | 55.2 (626) |
| Secondary+ | 20.5 (394) | 18.8 (147) | 21.8 (247) |
| Used net previous night, % (n) | 62.0 (1,190) | 64.4 (730) | 58.7 (460) |
| Improved walls, % (n) | 18.1 (348) | 18.0 (141) | 18.3 (348) |
| Improved floors, % (n) | 46.3 (888) | 49.8 (565) | 46.3 (888) |
| Household size, mean (SD) | 5.3 (2.1) | 5.7 (2.3) | 4.9 (1.8) |

.\* Villages were classified as high transmission if baseline prevalence of *P.falciparum* infection was > 10%.

### Travel among study participants

Participants were followed for an average of 10.3 months (Table 2). On average, 8.2% of participants reported any overnight travel over the prior two months at each follow-up, and over the full study period nearly one-third of participants reported traveling. Travel was more common among those over 15 years of age (11.8%) than among those under 5 (7.2%) and those 5-15 (3.4%), but only slightly more frequent among males (9.0%) vs females (6.7%). Trips were, on average, 1.7 weeks long with a wide range (1 - 62 days). The vast majority of travelers reported only one trip in the prior two months (97.0%). Nearly half of the trips were a short distance, within 25 kilometers of the village of residence (see Supplemental Figure 2 and Supplemental Table 1 for a map and table of travel destinations). 55% of trips were outside of the district but within Uganda, and only 0.9% were outside of the country. The most common reason for travel was visiting relatives or friends.

**Table 2.** Description of malaria and travel patterns among study participants, overall and stratified by village-level transmission intensity.

|  |  |  |  |
| --- | --- | --- | --- |
|  | All villages | High transmission villages | Low transmission villages |
| Number of participants, n | 1,918 | 784 | 1,134 |
| Number of follow-up months, mean (SD) | 10.3 (4.1) | 10.4 (4.2) | 10.2 (4.1) |
| Malaria episodes, n | 244 | 208 | 36 |
| Any self-reported overnight travel outside of village over study period, % | 29.4 | 32.3 | 27.3 |
| Any self-reported overnight travel outside of village in prior two months, % | 8.2 | 8.9 | 7.6 |
| Travel to higher incidence area in prior two months, % | 4.9 | 5.2 | 4.6 |
| Travel duration in weeks among travelers, mean (SD) | 1.7 (2.4) | 1.5 (2.3) | 2.2 (2.5) |
| Travel destination among travelers, % |  |  |  |
| Rukiga | 35.6 | 44.1 | 28.8 |
| Kampala | 14.4 | 14.2 | 14.6 |
| Rwanda | 21.5 | 16.6 | 25.5 |
| Other | 28.5 | 25.1 | 31.1 |
| Travel distance among trips, % |  |  |  |
| Less than 25 km | 47.8 | 53.0 | 43.6 |
| 25 to less than 50 km | 25.0 | 24.0 | 25.8 |
| Greater than 50 km | 27.2 | 23.0 | 30.6 |
| Travel reason among trips, % |  |  |  |
| Visiting relatives/friends | 65.7 | 69.3 | 62.8 |
| School | 9.2 | 4.2 | 13.3 |
| Work/trading | 10.3 | 10.8 | 9.9 |
| Other | 14.8 | 15.8 | 14.0 |

### Malaria burden

Over the study period, 244 episodes of malaria were recorded among cohort participants, 208 of which were among those residing in high transmission villages. There was substantial variation in malaria incidence over the study period, reflected in data from the health facility and in the cohort (Figure 2), with a peak in transmission in the first five months of the study followed by a period of sustained low transmission. In high transmission villages (where baseline parasite prevalence was at least 10%), median malaria incidence measured at the health facility was 331 cases per 1,000 person-years (PY, inter-quartile range [IQR] 96 to 536) in the first five study months, then declined nearly 10-fold (median 39 per 1,000 PY, IQR 17 to 60). In low transmission villages, median incidence in the first five study months was 41 cases per 1,000 PY (IQR 22 to 74) and declined to 12 per 1,000 PY (IQR 0 to 25). Similar trends were observed in parasite prevalence over time (Supplemental Figure 3).

**Figure 2.**
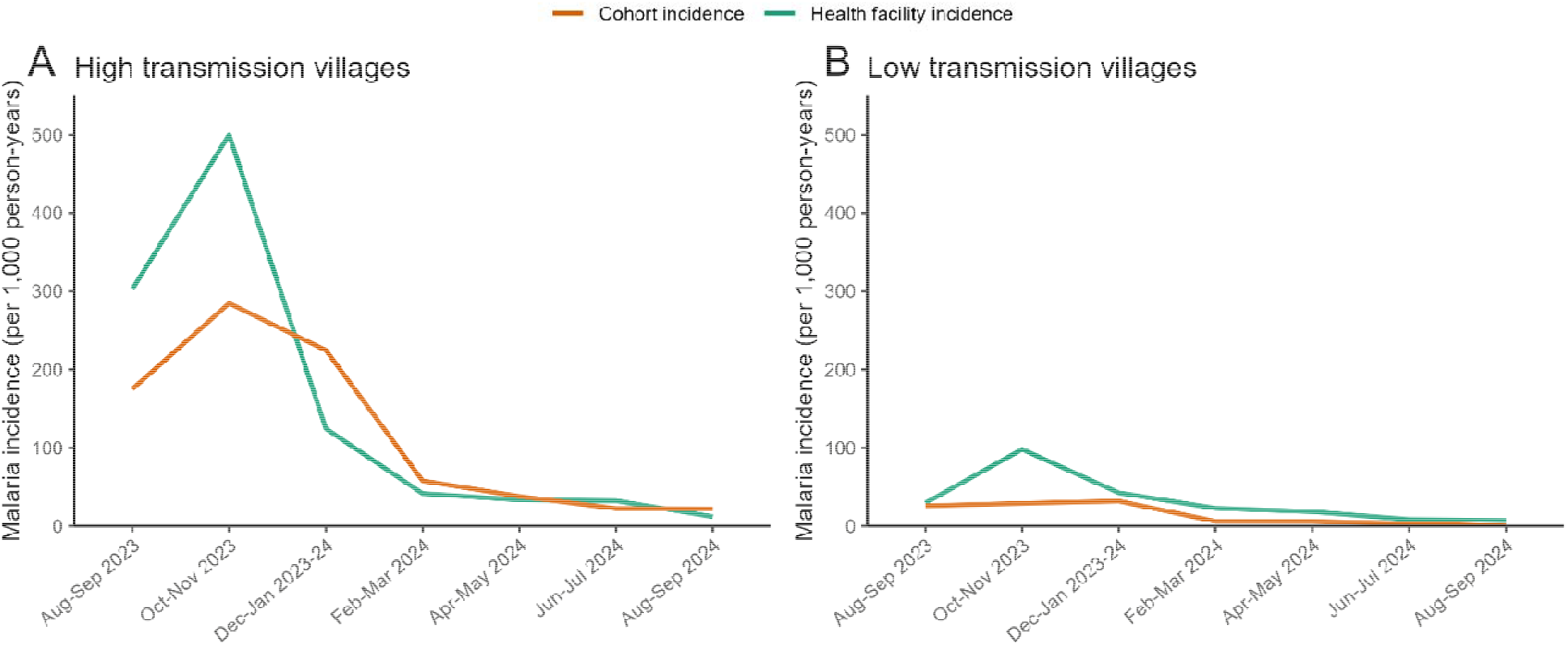
Malaria incidence over time measured through the cohort and at the health facility. Health facility-based incidence was calculated using cases from the study area that were captured at the facility as the numerator, and a population estimate from an enumeration survey as the denominator.

### Associations between overnight travel and malaria by transmission intensity

We first evaluated the relationship between any overnight travel and malaria. We found that overnight travel increased the odds of malaria by 86% (odds ratio [OR]=1.86, 95% CI 1.20-2.88); this relationship was stronger for residents in low transmission villages (OR=3.94, 95% CI 1.62-9.59) but no relationship was observed in high transmission villages (OR=1.30, 95% CI 0.80-2.11; Model 1 in Table 3, Supplemental Tables 2-6). Since travel-associated malaria may be more likely to occur when individuals travel to places with higher transmission intensity than the village where an individual resides, we then evaluated the association between overnight travel specifically to areas of higher incidence. Using this definition of travel, we found stronger associations across all villages (Model 2, OR=2.50, 95% CI 1.43-4.35), for those residing in low transmission villages (OR=5.65, 95% CI 2.06-15.45), and for those residing in high transmission villages (OR=1.86, 95% CI 1.06-3.25). These results suggest that travel to higher incidence areas may directly account for 6.8% of malaria cases overall and 17.8% of cases in low transmission villages. We also examined the role of trip duration. Among those in low transmission villages, short trips (<1 week) were not associated with malaria, but trips lasting one week or longer were associated with increased odds of malaria (OR=5.76, 95% CI 2.21-14.97) compared to no travel. Finally, we assessed the relationship between travel destination and malaria risk (Models 4 and 5). Trips within Rukiga district (where Kamwezi is located) were associated with increased malaria risk, while trips to other destinations were not. Consistent with this result, travel to destinations within 25 kilometers of the village was associated with greater odds of malaria, whereas longer-distance trips were not associated. Trips within Rukiga accounted for 4.3% of cases overall and 11.4% of cases in low transmission villages, suggesting that local travel accounted for approximately two-thirds of imported cases.

**Table 3.** Odds ratios from adjusted models assessing the association between travel variables and incident malaria among cohort members. Observations are from 1,918 participants from a total of 6 visits.

|  |  |  |  |  |
| --- | --- | --- | --- | --- |
|  |  | All villages | High transmission villages | Low transmission villages |
|  | Exposure | OR (95% CI) | OR (95% CI) | OR (95% CI) |
| Model 1 | Any overnight travel | 1.86* (1.20, 2.88) | 1.30 (0.80, 2.11) | 3.94** (1.62, 9.59) |
| Model 2 | Travel to higher incidence area | 2.50* (1.43, 4.35) | 1.86* (1.06, 3.25) | 5.65*** (2.06, 15.45) |
| Model 3 | Travel duration |  |  |  |
|  | No travel | REF | REF | REF |
|  | Less than one week | 1.86 (0.85, 4.08) | 1.55 (0.81, 2.98) | 1.31 (0.16, 10.88) |
|  | One week or greater | 1.86 (0.70, 4.96) | 1.06 (0.51, 2.18) | 5.76*** (2.21, 14.97) |
| Model 4 | Travel destination |  |  |  |
|  | No travel | REF | REF | REF |
|  | Rukiga | 4.06* (1.37, 12.03) | 2.94** (1.45, 5.94) | 10.72*** (3.20, 35.92) |
|  | Other | 1.54 (0.50, 4.77) | 1.08 (0.44, 2.62) | 3.84 (0.84, 17.50) |
| Model 5 | Travel distance |  |  |  |
|  | No travel | REF | REF | REF |
|  | Less than 25 km | 4.06 (0.94, 17.63) | 2.93** (1.45, 5.94) | 10.72*** (3.20, 35.96) |
|  | 25 to less than 50 km | 1.81 (0.29, 11.41) | 1.32 (0.34, 5.15) | 4.24 (0.56, 32.08) |
|  | 50 km and more | 1.42 (0.17, 11.95) | 0.97 (0.3, 3.08) | 3.66 (0.47, 28.34) |
| Adjusted for sex, age category, household improved floors, household improved walls, household size, and follow-up. |  |  |  |  |
| * <0.05, ** <0.01, *** <0.001 |  |  |  |  |

We repeated all analyses considering only laboratory-confirmed malaria cases diagnosed at the health facility for the outcome. These analyses included 8,255 observations of 1,816 individuals, with 111 incident malaria cases (two-thirds of the 186 cases captured during follow-ups that included self-reported cases). In these analyses, we found consistent associations between travel and malaria overall, but not in low transmission villages (Supplemental Tables 7 and 8). This may have been driven by the small number of cases captured in low transmission villages (n = 14).

To explore how our definition of “high” vs “low” transmission impacted the observed associations, we repeated the analyses exploring a range of thresholds and allowing the classification of villages to vary over time based on bi-monthly village-level parasite prevalence measured by qPCR (Figure 3). We found strong positive associations between travel and malaria under all definitions of low transmission, with the strength of the association increasing for lower thresholds. While overnight travel accounted for an estimated 10.5% of malaria in villages with parasite prevalence at or below 20%, it accounted for 29.3% of cases at villages with parasite prevalence below 5%. We also found moderate associations between overnight travel to higher incidence areas and malaria for villages classified as high transmission when using lower thresholds (i.e., those that included lower transmission villages), but not at thresholds at or above 30% prevalence.

**Figure 3.**
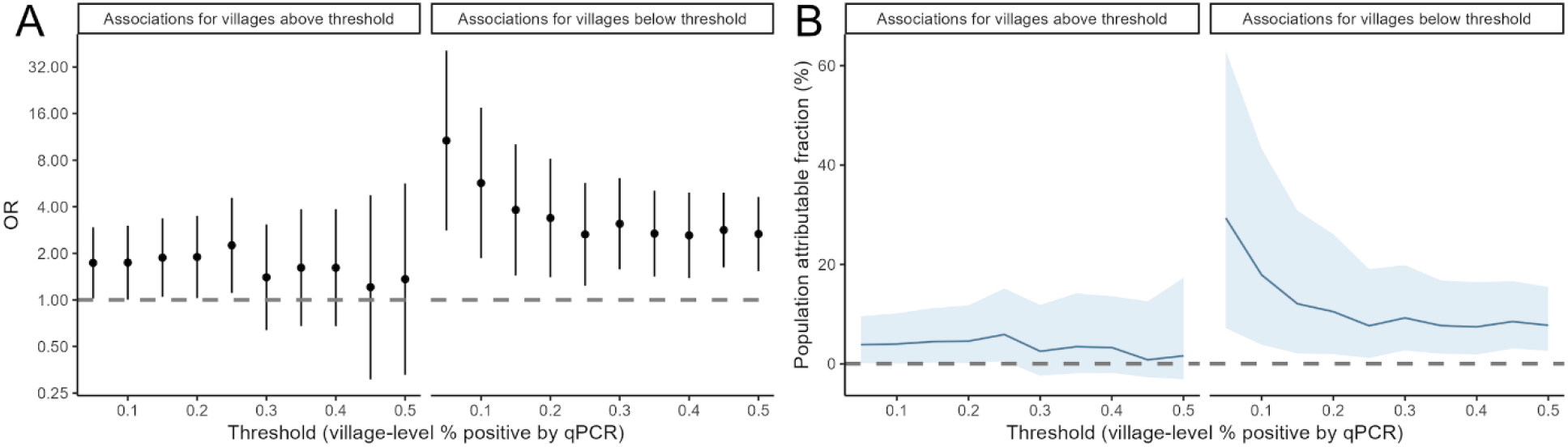
Relationship between travel to villages of higher incidence (ORs and 95% CI) and population attributable fractions when using varying thresholds to define high vs and low transmission villages.

### Associations between travel and malaria over time

Given the substantial decrease in malaria burden observed over time in the study area, we also assessed whether the associations between travel and malaria changed over time. When considering the full study area, we found that the association observed between travel to areas of higher transmission and malaria was mostly driven by strong positive associations during periods of lower local transmission (Figure 4). Furthermore, while travel was not associated with malaria incidence in high-transmission villages during high-transmission periods (before March 1, 2024), we found a strong association (OR=5.93, 95% CI 1.67-21.05) during low-transmission periods (from March 1, 2024, onward). This result implies that travel may directly account for at least 16.4% of malaria cases in high transmission villages during periods of low transmission. In low transmission villages travel was associated with malaria throughout the study duration; we estimate that it accounts for at least 11.6% of malaria cases during the high transmission period and 50.7% during the low transmission period. These results were consistent when considering only cases confirmed at the health facility as the outcome (Supplemental Figures 4 and 5).

**Figure 4.**
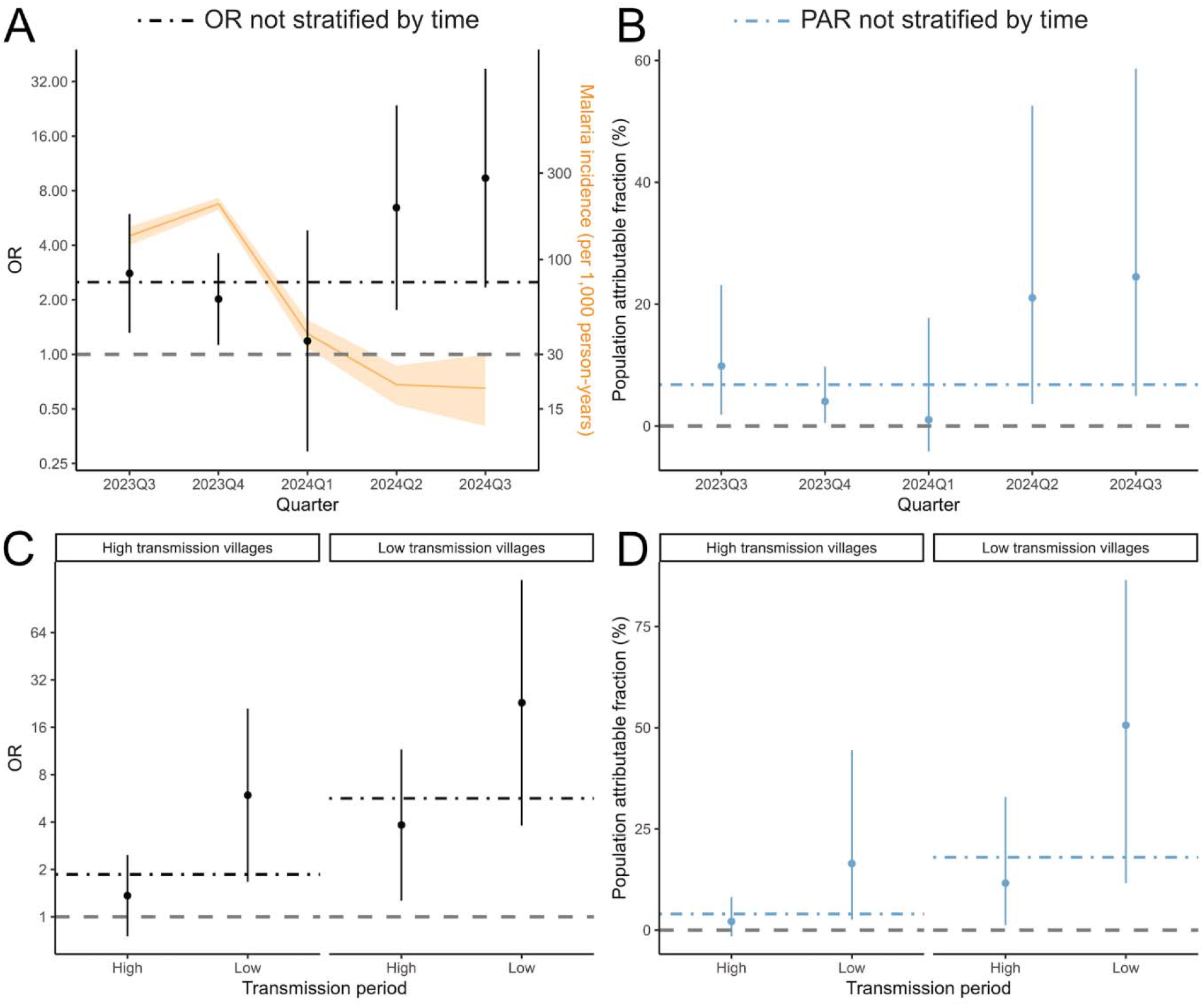
Relationship between travel to areas of higher incidence (ORs and 95% CI) and population attributable fractions over time and by time and village-level transmission intensity. Malaria incidence (and 95% CI) is shown in orange.

### Heterogeneity in malaria attributable to overnight travel over space and time

Finally, considering the variation in the associations between travel and malaria over space and time, we also estimated the attribution of travel on malaria cases for each village during high vs. low transmission periods (Figure 5). Over the full study period, we found that travel accounted for a median of 8.1% (IQR 3.9% to 16.5%) of malaria cases across all villages. In the high transmission period, we found that travel accounted for a median of 10.3% (IQR 7.8% to 13.6%) in low transmission villages; the confidence interval for PAFs in high transmission villages crossed zero. During the low transmission period, both high and low transmission villages demonstrated significant village-level PAFs (high transmission villages median 17.0%, IQR 13.3% to 20.4%; low transmission villages median 44.6%, IQR 37.1% to 51.8%) and travel accounted for up to 60% of malaria cases in some villages. PAFs by socio-demographic variables are presented in Supplemental Table 9.

**Figure 5.**
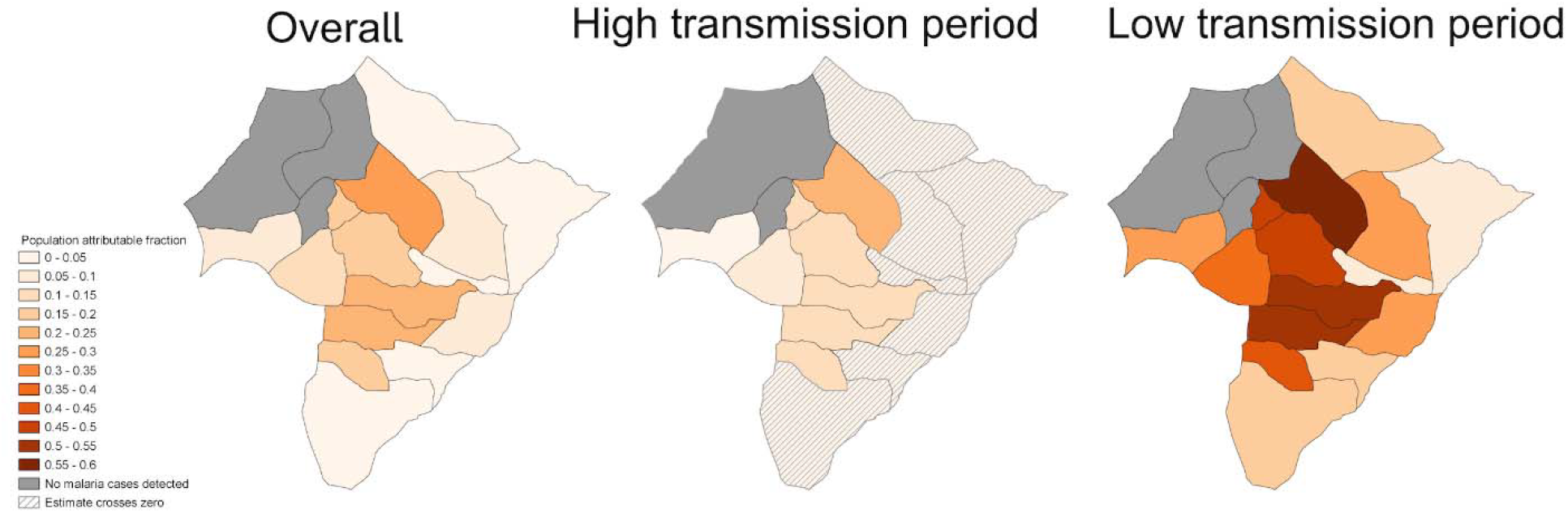
Village-level population attributable fractions representing the contribution of travel to higher incidence areas on malaria over the entire study period, during the high transmission period, and during the low transmission period.

## Discussion

We aimed to evaluate the role of importation on malaria in a low-to-moderate burden region of Uganda with substantial spatial and temporal heterogeneity in transmission intensity. We found that while the overall associations between travel and incident malaria were modest across the full study population, travel was strongly and consistently associated with malaria in low transmission villages and across all villages during periods of lower transmission. These findings suggest that travel to higher transmission areas contributes directly to malaria burden in places where local transmission does not fully account for continued cases, and that importation becomes increasingly consequential as local transmission declines even on small spatial and temporal scales.

In this study, the impact of importation varied considerably across both space and time. Previous studies on the impact of importation have primarily been limited to low transmission settings approaching elimination, with findings suggesting that in areas with near-zero local transmission, importation contributes to most (if not all) burden.^13-16^ Our study area experienced low-to-moderate incidence, with substantial heterogeneity across villages and over the course of the year. We observed that even within this context, at fine geographic and temporal scales, importation became more important when local exposure risk dropped. These findings align with a 2020 study in Zambia, which found that importation played a larger role in low transmission periods and origins, but diminished in influence when background transmission was high.^17^ Together, the evidence suggests that importation’s contribution to burden is dynamic and contingent on subtle shifts in local transmission context.

While we estimate that travel may directly account for up to 60% of malaria cases in some of the study villages, it is important to note that the estimates of population attributable risk derived from this study are likely to represent lower bounds of the true contribution of importation on malaria burden in this setting for two reasons. First, our estimates capture only the direct effect of overnight travel on malaria incidence, without accounting for subsequent secondary cases that may arise from imported infections. Future work will combine these findings with genomic parasite data to estimate transmission chains and quantify the impact of importation on downstream cases. Second, we do not account for additional potential sources of imported cases, including visitors^14,18^ and day trips. While these factors are harder to measure, they may also be key contributors to malaria cases in this setting.

There is a robust literature that suggests that travel is a risk factor for malaria across Africa.^19-23^ Our findings suggest that not only did travel increase malaria risk in this setting, but the characteristics of travel also mattered. Importantly, we found that local and regional mobility— rather than long-distance travel—was more predictive of risk and may account for two-thirds of imported cases in this setting. This has practical implications: malaria prevention efforts should not exclusively target international or cross-country travelers but also address the much more common pattern of local mobility. We also found that longer duration trips were associated with greater risk of importation, a finding reflected in other settings.^17,24^

While there has long been an emphasis on the importance of importation in low transmission, near-elimination settings^3^, our results indicate that it may be equally important in regions with moderate burden that have seen recent declines in local transmission, as illustrated by our high transmission villages. Notably, the most recent universal distribution of long-lasting insecticidal nets in the study area occurred in November 2023, coinciding with a marked decline in malaria incidence. These LLINs included those treated with chlorfenapyr and alpha-cypermethrin, which have demonstrated high efficacy.^25^ This timing raises the possibility that reduced local transmission following net distribution may have increased the relative contribution of travel to malaria burden. This finding has important public health implications. In these settings, National Malaria Control Programs may consider a hybrid approach, combining interventions that target both locally acquired infections (e.g., LLINs, IRS) and imported infections (e.g., active screening and treatment of travelers). Surveillance systems and prevention strategies should be designed to detect and mitigate travel-related importation, particularly in focal areas of low transmission and during periods of low transmission.

This study has several notable strengths. We drew on data from a large, well-characterized cohort with repeated measures over 14 months, enabling detailed temporal and spatial analyses. Rich travel histories allowed us to distinguish local from distant mobility and to detect short-distance importation patterns that are often overlooked. The combination of passive facility-based surveillance with active household follow-up enhanced case detection and improved attribution of malaria episodes. However, there are some limitations to this study. Our reliance on self-reported malaria and travel histories introduces potential for misclassification; however, we confirmed cases with health facility records when possible, and we do not expect reporting errors to differ by malaria status, suggesting any bias would likely be toward the null. Sensitivity analyses using confirmed cases were constrained by small sample sizes in low-transmission settings, potentially limiting power to detect associations. We also used external incidence estimates to classify destination transmission intensity, which may not fully capture local or seasonal variation. Finally, our study focused on symptomatic malaria cases, even though incidence of *P. falciparum* infection may be a more relevant outcome for quantifying the role of importation in sustaining transmission. As such, we may be underestimating the number of imported cases, but this should not bias our estimates importation’s contribution, unless the likelihood of asymptomatic infection differs between those who travel and those who don’t. Future studies could improve on these aspects by incorporating more granular spatial data, real-time case confirmation, and serological or genomic tools to validate exposure and infection.

Our findings indicate that the contribution of importation to malaria burden and transmission is dynamic and may be substantial even in areas that are not yet nearing elimination but are experiencing declines in local transmission. This underscores the need to move beyond a binary framework of control vs. elimination and consider adopting a more nuanced approach that recognizes the evolving role of importation across different transmission settings. Ministries of Health, including those in other high burden countries like Uganda, may consider implementing integrated strategies that address both locally acquired and imported infections, supported by responsive surveillance systems and adaptable intervention frameworks.

## Data Availability

The data used in this analysis will be made publicly available.

## Data sharing statement

The data used in this analysis will be made publicly available.

## Acknowledgements

This study was funded by the National Institute of Allergy and Infectious Diseases (NIAID; R01 AI163201). BG is funded by NIAID (K24 AI144048). IRB is a Chan Zuckerberg Biohub Investigator. We thank the study participants and the Kamwezi Health Centre staff and the Uganda Ministry of Health.

## Author contributions

AE and IRB conceived of the study, with input from GD, BG, and EA. IRB, AW, and EA designed the study protocol with help from IR. TMK, MO, and RT led fieldwork with oversight from EA. BAK, ST, and IW led the lab work with oversight from IS, JB, and MM. OA and IN led data management. AE performed data analyses with input from IRB. AE drafted the manuscript with IRB, GD, BG, and EA. All authors reviewed and edited the manuscript.

