## Supplemental tables and figures for "Quantifying the role of importation on sustained malaria transmission in a low-to-moderate burden region of Southwest Uganda"

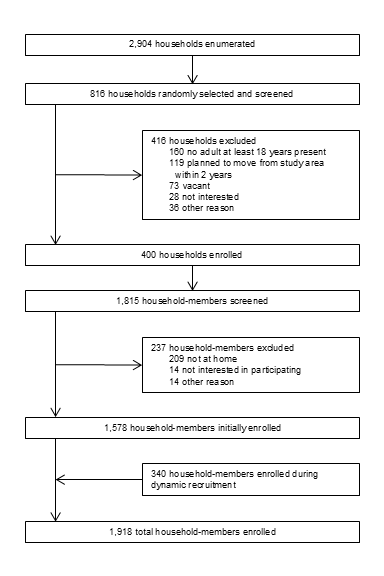

**Supplemental Figure 1.** Enrollment and follow-up of study participants.

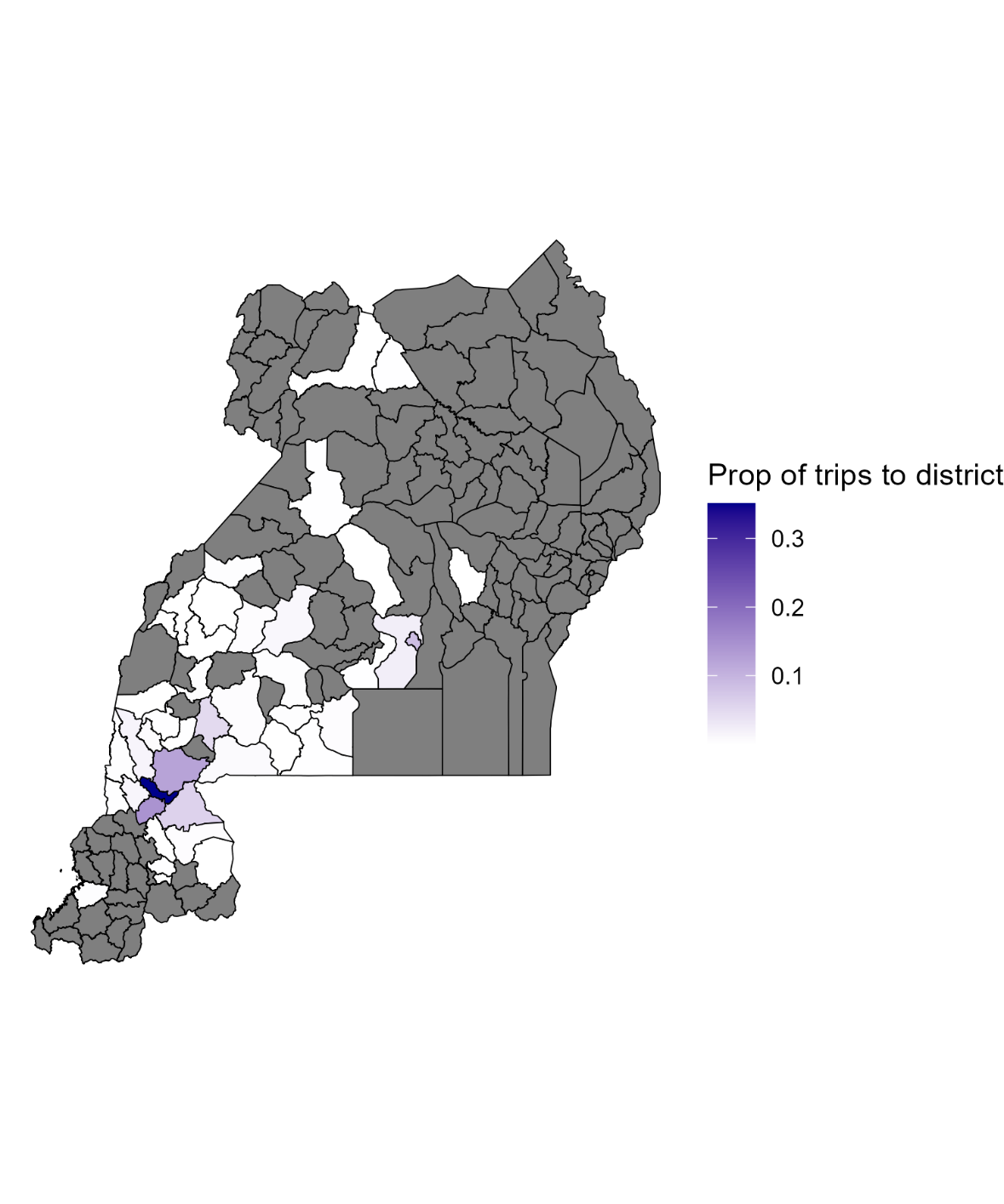

**Supplemental Figure 2**. Proportion of trips at the district-level among cohort members.

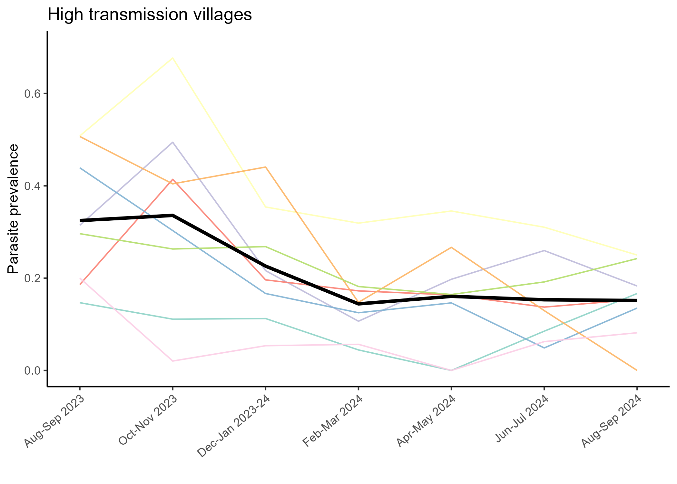

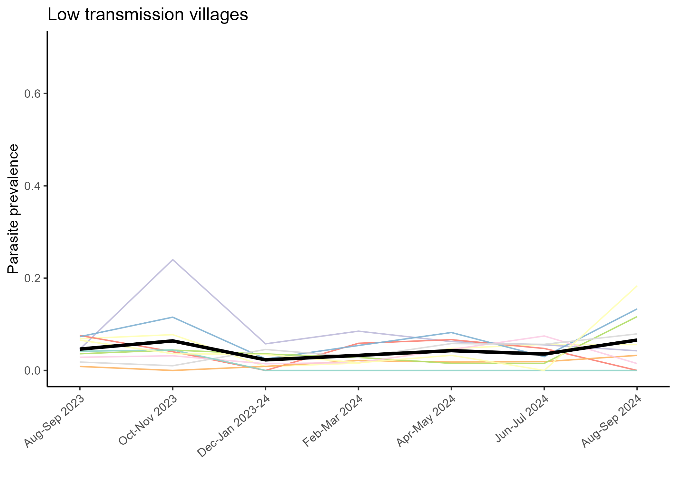

**Supplemental Figure 3.** Prevalence of *Plasmodium falciparum* over time stratified by village-level transmission intensity, presented as averages (black lines) and by village (colored lines).

| **Supplemental Table 1.** Destinations of trips among cohort members. | | | |
| --- | --- | --- | --- |
| District | Country | Proportion of trips | Count of trips |
| RUKIGA | Uganda | 0.351 | 254 |
| KABALE | Uganda | 0.145 | 105 |
| NTUNGAMO | Uganda | 0.123 | 89 |
| KAMPALA | Uganda | 0.087 | 63 |
| NYAGATARE | Rwanda | 0.059 | 43 |
| MBARARA | Uganda | 0.050 | 36 |
| WAKISO | Uganda | 0.024 | 17 |
| RUKUNGIRI | Uganda | 0.015 | 11 |
| RUBANDA | Uganda | 0.014 | 10 |
| MUBENDE | Uganda | 0.011 | 8 |
| ISINGIRO | Uganda | 0.008 | 6 |
| MITOOMA | Uganda | 0.007 | 5 |
| KIRUHURA | Uganda | 0.006 | 4 |
| KYEGEGWA | Uganda | 0.006 | 4 |
| MASAKA | Uganda | 0.006 | 4 |
| GATSIBO | Rwanda | 0.006 | 4 |
| IBANDA | Uganda | 0.004 | 3 |
| KAGADI | Uganda | 0.004 | 3 |
| KANUNGU | Uganda | 0.004 | 3 |
| LWENGO | Uganda | 0.004 | 3 |
| SSEMBABULE | Uganda | 0.004 | 3 |
| KARONGI | Rwanda | 0.004 | 3 |
| BUSHENYI | Uganda | 0.003 | 2 |
| GULU | Uganda | 0.003 | 2 |
| KISORO | Uganda | 0.003 | 2 |
| KYOTERA | Uganda | 0.003 | 2 |
| MASINDI | Uganda | 0.003 | 2 |
| MPIGI | Uganda | 0.003 | 2 |
| GASABO | Rwanda | 0.003 | 2 |
| SHEEMA | Uganda | 0.003 | 2 |
| AMURU | Uganda | 0.001 | 1 |
| BUNYANGABU | Uganda | 0.001 | 1 |
| KABAROLE | Uganda | 0.001 | 1 |
| KAMULI | Uganda | 0.001 | 1 |
| KAMWENGE | Uganda | 0.001 | 1 |
| KYENJOJO | Uganda | 0.001 | 1 |
| NAKASEKE | Uganda | 0.001 | 1 |
| RAKAI | Uganda | 0.001 | 1 |
| KICUKIRO | Rwanda | 0.001 | 1 |
| RUBIRIZI | Uganda | 0.001 | 1 |
| KAYONZA | Rwanda | 0.001 | 1 |
| GICUMBI | Rwanda | 0.001 | 1 |

| **Supplemental Table 2.** Odds ratios from adjusted models assessing the association between travel and incident malaria among cohort members. | | | | | | | | | |
| --- | --- | --- | --- | --- | --- | --- | --- | --- | --- |
|  | **All villages** | | | **High transmission villages** | | | **Low transmission villages** | | |
| **Characteristic** | **OR***^1^* | **95% CI***^1^* | **p-value** | **OR***^1^* | **95% CI***^1^* | **p-value** | **OR***^1^* | **95% CI***^1^* | **p-value** |
| Any overnight travel |  |  |  |  |  |  |  |  |  |
| No travel | — | — |  | — | — |  | — | — |  |
| Travel | 1.86 | 1.20, 2.88 | 0.017 | 1.30 | 0.80, 2.11 | 0.3 | 3.94 | 1.62, 9.59 | 0.003 |
| Male gender | 0.96 | 0.68, 1.34 | 0.7 | 1.01 | 0.72, 1.43 | >0.9 | 1.22 | 0.61, 2.47 | 0.6 |
| Age category |  |  |  |  |  |  |  |  |  |
| < 5 | — | — |  | — | — |  | — | — |  |
| 5-15 | 1.53 | 0.81, 2.89 | 0.13 | 1.39 | 0.88, 2.17 | 0.2 | 4.77 | 1.08, 21.0 | 0.039 |
| > 15 | 0.79 | 0.34, 1.84 | 0.5 | 0.79 | 0.49, 1.25 | 0.3 | 1.91 | 0.43, 8.51 | 0.4 |
| Improved walls | 0.65 | 0.24, 1.74 | 0.3 | 0.65 | 0.36, 1.17 | 0.15 | 0.51 | 0.14, 1.87 | 0.3 |
| Improved floor | 0.55 | 0.30, 1.01 | 0.053 | 0.59 | 0.39, 0.90 | 0.016 | 1.16 | 0.55, 2.47 | 0.7 |
| Household size | 1.16 | 0.96, 1.39 | 0.093 | 1.09 | 1.00, 1.18 | 0.048 | 0.93 | 0.74, 1.17 | 0.5 |
| Follow-up |  |  |  |  |  |  |  |  |  |
| 0 | — | — |  | — | — |  | — | — |  |
| 1 | 1.65 | 0.92, 2.96 | 0.076 | 1.71 | 1.18, 2.47 | 0.004 | 1.24 | 0.53, 2.89 | 0.6 |
| 2 | 1.29 | 0.73, 2.31 | 0.3 | 1.30 | 0.87, 1.94 | 0.2 | 1.26 | 0.53, 3.01 | 0.6 |
| 3 | 0.31 | 0.09, 1.08 | 0.060 | 0.31 | 0.17, 0.56 | <0.001 | 0.23 | 0.05, 1.05 | 0.057 |
| 4 | 0.21 | 0.09, 0.51 | 0.008 | 0.20 | 0.10, 0.42 | <0.001 | 0.23 | 0.05, 1.05 | 0.058 |
| 5 | 0.12 | 0.02, 0.66 | 0.026 | 0.12 | 0.05, 0.30 | <0.001 | 0.12 | 0.02, 0.96 | 0.045 |
| 6 | 0.10 | 0.04, 0.30 | 0.004 | 0.11 | 0.05, 0.29 | <0.001 | 0.00 | 0.00, 0.00 | <0.001 |
| *^1^*OR = Odds Ratio, CI = Confidence Interval | | | | | | | | | |

| **Supplemental Table 3.** Odds ratios from adjusted models assessing the association between travel to an area of higher incidence and incident malaria among cohort members. | | | | | | | | | |
| --- | --- | --- | --- | --- | --- | --- | --- | --- | --- |
|  | **All villages** | | | **High transmission villages** | | | **Low transmission villages** | | |
| **Characteristic** | **OR***^1^* | **95% CI***^1^* | **p-value** | **OR***^1^* | **95% CI***^1^* | **p-value** | **OR***^1^* | **95% CI***^1^* | **p-value** |
| Travel to higher incidence area |  |  |  |  |  |  |  |  |  |
| No travel | — | — |  | — | — |  | — | — |  |
| Travel | 2.50 | 1.43, 4.35 | 0.010 | 1.86 | 1.06, 3.25 | 0.029 | 5.65 | 2.06, 15.4 | <0.001 |
| Male gender | 0.95 | 0.68, 1.34 | 0.7 | 1.01 | 0.72, 1.42 | >0.9 | 1.20 | 0.59, 2.45 | 0.6 |
| Age category |  |  |  |  |  |  |  |  |  |
| < 5 | — | — |  | — | — |  | — | — |  |
| 5-15 | 1.53 | 0.81, 2.89 | 0.14 | 1.40 | 0.89, 2.18 | 0.14 | 4.86 | 1.09, 21.7 | 0.038 |
| > 15 | 0.78 | 0.33, 1.83 | 0.5 | 0.77 | 0.48, 1.23 | 0.3 | 1.98 | 0.44, 8.95 | 0.4 |
| Improved walls | 0.64 | 0.24, 1.71 | 0.3 | 0.65 | 0.36, 1.16 | 0.15 | 0.49 | 0.13, 1.75 | 0.3 |
| Improved floor | 0.56 | 0.31, 1.02 | 0.055 | 0.59 | 0.38, 0.90 | 0.014 | 1.23 | 0.57, 2.70 | 0.6 |
| Household size | 1.16 | 0.96, 1.39 | 0.10 | 1.09 | 1.00, 1.18 | 0.046 | 0.93 | 0.74, 1.17 | 0.5 |
| Follow-up |  |  |  |  |  |  |  |  |  |
| 0 | — | — |  | — | — |  | — | — |  |
| 1 | 1.65 | 0.92, 2.97 | 0.077 | 1.70 | 1.18, 2.46 | 0.005 | 1.26 | 0.53, 2.97 | 0.6 |
| 2 | 1.30 | 0.72, 2.32 | 0.3 | 1.30 | 0.87, 1.94 | 0.2 | 1.29 | 0.54, 3.09 | 0.6 |
| 3 | 0.30 | 0.09, 1.08 | 0.059 | 0.30 | 0.17, 0.55 | <0.001 | 0.23 | 0.05, 1.05 | 0.057 |
| 4 | 0.21 | 0.09, 0.51 | 0.008 | 0.20 | 0.10, 0.41 | <0.001 | 0.23 | 0.05, 1.08 | 0.063 |
| 5 | 0.12 | 0.02, 0.66 | 0.026 | 0.12 | 0.05, 0.30 | <0.001 | 0.12 | 0.02, 0.98 | 0.047 |
| 6 | 0.10 | 0.04, 0.30 | 0.004 | 0.12 | 0.05, 0.29 | <0.001 | 0.00 | 0.00, 0.00 | <0.001 |
| *^1^*OR = Odds Ratio, CI = Confidence Interval | | | | | | | | | |

| **Supplemental Table 4.** Odds ratios from adjusted models assessing the association between travel duration and incident malaria among cohort members. | | | | | | | | | |
| --- | --- | --- | --- | --- | --- | --- | --- | --- | --- |
|  | **All villages** | | | **High transmission villages** | | | **Low transmission villages** | | |
| **Characteristic** | **OR***^1^* | **95% CI***^1^* | **p-value** | **OR***^1^* | **95% CI***^1^* | **p-value** | **OR***^1^* | **95% CI***^1^* | **p-value** |
| Travel duration |  |  |  |  |  |  |  |  |  |
| No travel | — | — |  | — | — |  | — | — |  |
| Less than 1 week | 1.86 | 0.85, 4.08 | 0.087 | 1.55 | 0.81, 2.98 | 0.2 | 1.31 | 0.16, 10.9 | 0.8 |
| 1 week or greater | 1.86 | 0.70, 4.96 | 0.14 | 1.06 | 0.51, 2.18 | 0.9 | 5.76 | 2.21, 15.0 | <0.001 |
| Male gender | 0.96 | 0.65, 1.41 | 0.8 | 1.02 | 0.72, 1.43 | >0.9 | 1.22 | 0.61, 2.46 | 0.6 |
| Age category |  |  |  |  |  |  |  |  |  |
| < 5 | — | — |  | — | — |  | — | — |  |
| 5-15 | 1.53 | 0.74, 3.18 | 0.2 | 1.39 | 0.89, 2.18 | 0.15 | 4.67 | 1.06, 20.6 | 0.041 |
| > 15 | 0.79 | 0.30, 2.07 | 0.5 | 0.78 | 0.49, 1.25 | 0.3 | 1.99 | 0.44, 8.94 | 0.4 |
| Improved walls | 0.65 | 0.21, 2.01 | 0.3 | 0.65 | 0.36, 1.17 | 0.15 | 0.51 | 0.14, 1.83 | 0.3 |
| Improved floor | 0.55 | 0.28, 1.11 | 0.073 | 0.59 | 0.39, 0.91 | 0.016 | 1.13 | 0.54, 2.39 | 0.7 |
| Household size | 1.16 | 0.94, 1.43 | 0.11 | 1.08 | 1.00, 1.18 | 0.050 | 0.93 | 0.74, 1.17 | 0.5 |
| Follow-up |  |  |  |  |  |  |  |  |  |
| 0 | — | — |  | — | — |  | — | — |  |
| 1 | 1.65 | 0.85, 3.21 | 0.10 | 1.71 | 1.18, 2.47 | 0.004 | 1.19 | 0.51, 2.80 | 0.7 |
| 2 | 1.29 | 0.67, 2.52 | 0.3 | 1.30 | 0.87, 1.94 | 0.2 | 1.18 | 0.48, 2.85 | 0.7 |
| 3 | 0.31 | 0.07, 1.30 | 0.080 | 0.30 | 0.17, 0.55 | <0.001 | 0.22 | 0.05, 1.00 | 0.050 |
| 4 | 0.21 | 0.08, 0.58 | 0.016 | 0.20 | 0.10, 0.41 | <0.001 | 0.22 | 0.05, 0.98 | 0.047 |
| 5 | 0.12 | 0.02, 0.85 | 0.041 | 0.12 | 0.05, 0.30 | <0.001 | 0.12 | 0.01, 0.90 | 0.040 |
| 6 | 0.10 | 0.03, 0.35 | 0.010 | 0.11 | 0.05, 0.29 | <0.001 | 0.00 | 0.00, 0.00 | <0.001 |
| *^1^*OR = Odds Ratio, CI = Confidence Interval | | | | | | | | | |

| **Supplemental Table 5.** Odds ratios from adjusted models assessing the association between travel destination and incident malaria among cohort members. | | | | | | | | | |
| --- | --- | --- | --- | --- | --- | --- | --- | --- | --- |
|  | **All villages** | | | **High transmission villages** | | | **Low transmission villages** | | |
| **Characteristic** | **OR***^1^* | **95% CI***^1^* | **p-value** | **OR***^1^* | **95% CI***^1^* | **p-value** | **OR***^1^* | **95% CI***^1^* | **p-value** |
| Travel destination |  |  |  |  |  |  |  |  |  |
| No travel | — | — |  | — | — |  | — | — |  |
| RUKIGA | 4.06 | 1.37, 12.0 | 0.026 | 2.94 | 1.45, 5.94 | 0.003 | 10.7 | 3.20, 35.9 | <0.001 |
| Other | 1.54 | 0.50, 4.77 | 0.3 | 1.08 | 0.44, 2.62 | 0.9 | 3.84 | 0.84, 17.5 | 0.082 |
| Male gender | 0.96 | 0.65, 1.42 | 0.8 | 1.02 | 0.72, 1.44 | >0.9 | 1.21 | 0.59, 2.48 | 0.6 |
| Age category |  |  |  |  |  |  |  |  |  |
| < 5 | — | — |  | — | — |  | — | — |  |
| 5-15 | 1.54 | 0.74, 3.21 | 0.2 | 1.39 | 0.89, 2.18 | 0.15 | 5.09 | 1.10, 23.5 | 0.037 |
| > 15 | 0.78 | 0.29, 2.10 | 0.5 | 0.77 | 0.48, 1.23 | 0.3 | 2.09 | 0.46, 9.57 | 0.3 |
| Improved walls | 0.64 | 0.21, 1.97 | 0.3 | 0.65 | 0.36, 1.17 | 0.2 | 0.47 | 0.14, 1.62 | 0.2 |
| Improved floor | 0.56 | 0.28, 1.13 | 0.078 | 0.59 | 0.38, 0.90 | 0.015 | 1.28 | 0.59, 2.79 | 0.5 |
| Household size | 1.15 | 0.93, 1.42 | 0.12 | 1.08 | 1.00, 1.17 | 0.052 | 0.93 | 0.74, 1.17 | 0.5 |
| Follow-up |  |  |  |  |  |  |  |  |  |
| 0 | — | — |  | — | — |  | — | — |  |
| 1 | 1.64 | 0.83, 3.23 | 0.10 | 1.69 | 1.17, 2.45 | 0.005 | 1.26 | 0.53, 2.98 | 0.6 |
| 2 | 1.31 | 0.67, 2.57 | 0.3 | 1.31 | 0.88, 1.95 | 0.2 | 1.34 | 0.56, 3.22 | 0.5 |
| 3 | 0.30 | 0.07, 1.30 | 0.080 | 0.30 | 0.17, 0.55 | <0.001 | 0.23 | 0.05, 1.06 | 0.059 |
| 4 | 0.21 | 0.08, 0.57 | 0.016 | 0.20 | 0.10, 0.41 | <0.001 | 0.23 | 0.05, 1.08 | 0.063 |
| 5 | 0.12 | 0.02, 0.86 | 0.042 | 0.12 | 0.05, 0.31 | <0.001 | 0.13 | 0.02, 1.03 | 0.053 |
| 6 | 0.10 | 0.03, 0.35 | 0.009 | 0.11 | 0.05, 0.28 | <0.001 | 0.00 | 0.00, 0.00 | <0.001 |
| *^1^*OR = Odds Ratio, CI = Confidence Interval | | | | | | | | | |

| **Supplemental Table 6.** Odds ratios from adjusted models assessing the association between travel distance and incident malaria among cohort members. | | | | | | | | | |
| --- | --- | --- | --- | --- | --- | --- | --- | --- | --- |
|  | **All villages** | | | **High transmission villages** | | | **Low transmission villages** | | |
| **Characteristic** | **OR***^1^* | **95% CI***^1^* | **p-value** | **OR***^1^* | **95% CI***^1^* | **p-value** | **OR***^1^* | **95% CI***^1^* | **p-value** |
| Travel distance |  |  |  |  |  |  |  |  |  |
| No travel | — | — |  | — | — |  | — | — |  |
| Less than 25 km | 4.06 | 0.94, 17.6 | 0.054 | 2.93 | 1.45, 5.94 | 0.003 | 10.7 | 3.20, 36.0 | <0.001 |
| 25- <50km | 1.81 | 0.29, 11.4 | 0.3 | 1.32 | 0.34, 5.15 | 0.7 | 4.24 | 0.56, 32.1 | 0.2 |
| 50km + | 1.42 | 0.17, 12.0 | 0.6 | 0.97 | 0.30, 3.08 | >0.9 | 3.66 | 0.47, 28.3 | 0.2 |
| Male gender | 0.96 | 0.56, 1.64 | 0.8 | 1.02 | 0.72, 1.44 | >0.9 | 1.21 | 0.59, 2.48 | 0.6 |
| Age category |  |  |  |  |  |  |  |  |  |
| < 5 | — | — |  | — | — |  | — | — |  |
| 5-15 | 1.55 | 0.57, 4.19 | 0.2 | 1.40 | 0.89, 2.19 | 0.14 | 5.10 | 1.10, 23.6 | 0.037 |
| > 15 | 0.79 | 0.21, 2.99 | 0.5 | 0.77 | 0.48, 1.23 | 0.3 | 2.09 | 0.46, 9.59 | 0.3 |
| Improved walls | 0.64 | 0.14, 2.93 | 0.3 | 0.65 | 0.36, 1.17 | 0.2 | 0.47 | 0.14, 1.64 | 0.2 |
| Improved floor | 0.56 | 0.22, 1.44 | 0.12 | 0.59 | 0.38, 0.91 | 0.016 | 1.28 | 0.59, 2.78 | 0.5 |
| Household size | 1.15 | 0.87, 1.53 | 0.2 | 1.08 | 1.00, 1.17 | 0.051 | 0.93 | 0.74, 1.17 | 0.5 |
| Follow-up |  |  |  |  |  |  |  |  |  |
| 0 | — | — |  | — | — |  | — | — |  |
| 1 | 1.64 | 0.65, 4.11 | 0.15 | 1.69 | 1.17, 2.45 | 0.005 | 1.26 | 0.54, 2.97 | 0.6 |
| 2 | 1.31 | 0.53, 3.26 | 0.3 | 1.31 | 0.88, 1.95 | 0.2 | 1.35 | 0.57, 3.21 | 0.5 |
| 3 | 0.31 | 0.04, 2.16 | 0.12 | 0.30 | 0.17, 0.55 | <0.001 | 0.23 | 0.05, 1.04 | 0.056 |
| 4 | 0.21 | 0.05, 0.81 | 0.038 | 0.20 | 0.10, 0.41 | <0.001 | 0.23 | 0.05, 1.08 | 0.063 |
| 5 | 0.12 | 0.01, 1.71 | 0.076 | 0.12 | 0.05, 0.31 | <0.001 | 0.13 | 0.02, 1.03 | 0.053 |
| 6 | 0.10 | 0.02, 0.53 | 0.027 | 0.11 | 0.05, 0.28 | <0.001 | 0.00 | 0.00, 0.00 | <0.001 |
| *^1^*OR = Odds Ratio, CI = Confidence Interval | | | | | | | | | |

| **Supplemental Table 7.** Odds ratios from adjusted models assessing the association between travel and incident malaria among cohort members, with only confirmed malaria cases as outcome. | | | | | | | | | |
| --- | --- | --- | --- | --- | --- | --- | --- | --- | --- |
|  | **All villages** | | | **High transmission villages** | | | **Low transmission villages** | | |
| **Characteristic** | **OR***^1^* | **95% CI***^1^* | **p-value** | **OR***^1^* | **95% CI***^1^* | **p-value** | **OR***^1^* | **95% CI***^1^* | **p-value** |
| Any overnight travel |  |  |  |  |  |  |  |  |  |
| No travel | — | — |  | — | — |  | — | — |  |
| Travel | 2.13 | 1.30, 3.47 | 0.011 | 1.80 | 0.92, 3.56 | 0.088 | 2.74 | 0.44, 17.0 | 0.3 |
| Male gender | 0.89 | 0.60, 1.34 | 0.5 | 0.83 | 0.50, 1.37 | 0.5 | 2.63 | 0.79, 8.70 | 0.11 |
| Age category |  |  |  |  |  |  |  |  |  |
| < 5 | — | — |  | — | — |  | — | — |  |
| 5-15 | 2.36 | 0.85, 6.61 | 0.084 | 2.50 | 1.18, 5.28 | 0.017 | 2.76 | 0.34, 22.2 | 0.3 |
| > 15 | 1.27 | 0.52, 3.08 | 0.5 | 1.28 | 0.60, 2.76 | 0.5 | 2.37 | 0.28, 19.7 | 0.4 |
| Improved walls | 0.60 | 0.15, 2.48 | 0.4 | 0.57 | 0.22, 1.53 | 0.3 | 0.38 | 0.04, 3.53 | 0.4 |
| Improved floor | 0.44 | 0.18, 1.05 | 0.059 | 0.49 | 0.26, 0.92 | 0.025 | 0.87 | 0.26, 2.95 | 0.8 |
| Household size | 1.31 | 0.97, 1.76 | 0.070 | 1.23 | 1.08, 1.39 | 0.002 | 0.96 | 0.73, 1.25 | 0.7 |
| Follow-up |  |  |  |  |  |  |  |  |  |
| 0 | — | — |  | — | — |  | — | — |  |
| 1 | 2.02 | 1.01, 4.07 | 0.049 | 2.11 | 1.32, 3.36 | 0.002 | 2.23 | 0.66, 7.53 | 0.2 |
| 2 | 0.59 | 0.15, 2.38 | 0.4 | 0.58 | 0.31, 1.11 | 0.10 | 0.64 | 0.11, 3.58 | 0.6 |
| 3 | 0.30 | 0.12, 0.74 | 0.019 | 0.25 | 0.10, 0.63 | 0.003 | 0.64 | 0.12, 3.53 | 0.6 |
| 4 | 0.16 | 0.02, 1.51 | 0.089 | 0.18 | 0.06, 0.54 | 0.002 | 0.00 | 0.00, 0.00 | <0.001 |
| 5 | 0.12 | 0.03, 0.48 | 0.011 | 0.13 | 0.04, 0.44 | 0.001 | 0.00 | 0.00, 0.00 | <0.001 |
| 6 | 2.13 | 1.30, 3.47 | 0.011 | 1.80 | 0.92, 3.56 | 0.088 | 2.74 | 0.44, 17.0 | 0.3 |
| *^1^*OR = Odds Ratio, CI = Confidence Interval | | | | | | | | | |

| **Supplemental Table 8.** Odds ratios from adjusted models assessing the association between travel to an area of higher incidence and incident malaria among cohort members, with only confirmed malaria cases as outcome. | | | | | | | | | |
| --- | --- | --- | --- | --- | --- | --- | --- | --- | --- |
|  | **All villages** | | | **High transmission villages** | | | **Low transmission villages** | | |
| **Characteristic** | **OR***^1^* | **95% CI***^1^* | **p-value** | **OR***^1^* | **95% CI***^1^* | **p-value** | **OR***^1^* | **95% CI***^1^* | **p-value** |
| Travel to higher incidence area |  |  |  |  |  |  |  |  |  |
| No travel | — | — |  | — | — |  | — | — |  |
| Travel | 2.84 | 1.49, 5.42 | 0.009 | 2.43 | 1.13, 5.24 | 0.024 | 4.97 | 0.74, 33.2 | 0.10 |
| Male gender | 0.89 | 0.59, 1.34 | 0.5 | 0.82 | 0.50, 1.36 | 0.4 | 2.68 | 0.81, 8.85 | 0.10 |
| Age category |  |  |  |  |  |  |  |  |  |
| < 5 | — | — |  | — | — |  | — | — |  |
| 5-15 | 2.36 | 0.84, 6.67 | 0.086 | 2.49 | 1.18, 5.25 | 0.017 | 2.94 | 0.36, 24.3 | 0.3 |
| > 15 | 1.27 | 0.53, 3.01 | 0.5 | 1.27 | 0.59, 2.73 | 0.5 | 2.52 | 0.29, 21.6 | 0.4 |
| Improved walls | 0.60 | 0.15, 2.44 | 0.4 | 0.58 | 0.22, 1.55 | 0.3 | 0.37 | 0.04, 3.49 | 0.4 |
| Improved floor | 0.44 | 0.18, 1.06 | 0.061 | 0.48 | 0.26, 0.91 | 0.025 | 0.88 | 0.25, 3.09 | 0.8 |
| Household size | 1.30 | 0.97, 1.76 | 0.072 | 1.22 | 1.08, 1.39 | 0.002 | 0.95 | 0.73, 1.25 | 0.7 |
| Follow-up |  |  |  |  |  |  |  |  |  |
| 0 | — | — |  | — | — |  | — | — |  |
| 1 | 2.04 | 1.00, 4.13 | 0.049 | 2.13 | 1.33, 3.39 | 0.002 | 2.18 | 0.64, 7.38 | 0.2 |
| 2 | 0.59 | 0.14, 2.39 | 0.4 | 0.58 | 0.30, 1.10 | 0.10 | 0.61 | 0.11, 3.28 | 0.6 |
| 3 | 0.29 | 0.12, 0.72 | 0.017 | 0.25 | 0.10, 0.63 | 0.003 | 0.63 | 0.11, 3.45 | 0.6 |
| 4 | 0.16 | 0.02, 1.54 | 0.092 | 0.18 | 0.06, 0.54 | 0.002 | 0.00 | 0.00, 0.00 | <0.001 |
| 5 | 0.12 | 0.03, 0.48 | 0.011 | 0.13 | 0.04, 0.45 | 0.001 | 0.00 | 0.00, 0.00 | <0.001 |
| 6 | 2.84 | 1.49, 5.42 | 0.009 | 2.43 | 1.13, 5.24 | 0.024 | 4.97 | 0.74, 33.2 | 0.10 |
| *^1^*OR = Odds Ratio, CI = Confidence Interval | | | | | | | | | |

| **Supplemental Table 9.** Population attributable fractions (percentages) representing the contribution of travel to higher incidence areas on malaria by demographics, overall and by transmission season. | | | |
| --- | --- | --- | --- |
|  | All villages PAF (95% CI) | High transmission villages PAF (95% CI) | Low transmission villages PAF (95% CI) |
| Sex | | | |
| Male | 6.8 (2, 14) | 4 (0.3, 9.8) | 18.4 (4.9, 41.2) |
| Female | 3 (0.9, 6.5) | 1.8 (0.1, 4.5) | 8.8 (2.2, 23.2) |
| Age, % (n) | | | |
| <5 | 9.3 (2.9, 18.7) | 5.6 (0.4, 13.4) | 24.2 (6.8, 49.8) |
| 5-15 | 7.2 (2.2, 14.7) | 4.3 (0.3, 10.4) | 19.4 (5.2, 42.7) |
| >15 | 6.1 (1.8, 12.7) | 3.6 (0.3, 8.9) | 16.8 (4.4, 38.6) |
| Occupation | | | |
| Commerce | 8.3 (2.5, 16.9) | 5 (0.4, 12) | 22 (6, 46.7) |
| Farmer | 8.7 (2.7, 17.6) | 5.2 (0.4, 12.5) | 22.9 (6.3, 47.9) |
| Student | 3.5 (1, 7.4) | 2 (0.1, 5.1) | 10 (2.5, 25.7) |
| Childcare/homemaker | 8.4 (2.6, 17) | 5 (0.4, 12.1) | 22.1 (6.1, 46.8) |
| Other | 16 (5.2, 29.8) | 9.8 (0.8, 22.2) | 37.1 (11.9, 64.7) |
| Education level | | | |
| None | 6.8 (2.1, 14.1) | 4 (0.3, 9.9) | 18.5 (4.9, 41.4) |
| Primary | 5.4 (1.6, 11.3) | 3.2 (0.2, 7.9) | 15 (3.9, 35.4) |
| Secondary+ | 9.9 (3.1, 19.7) | 5.9 (0.4, 14.1) | 25.4 (7.2, 51.4) |

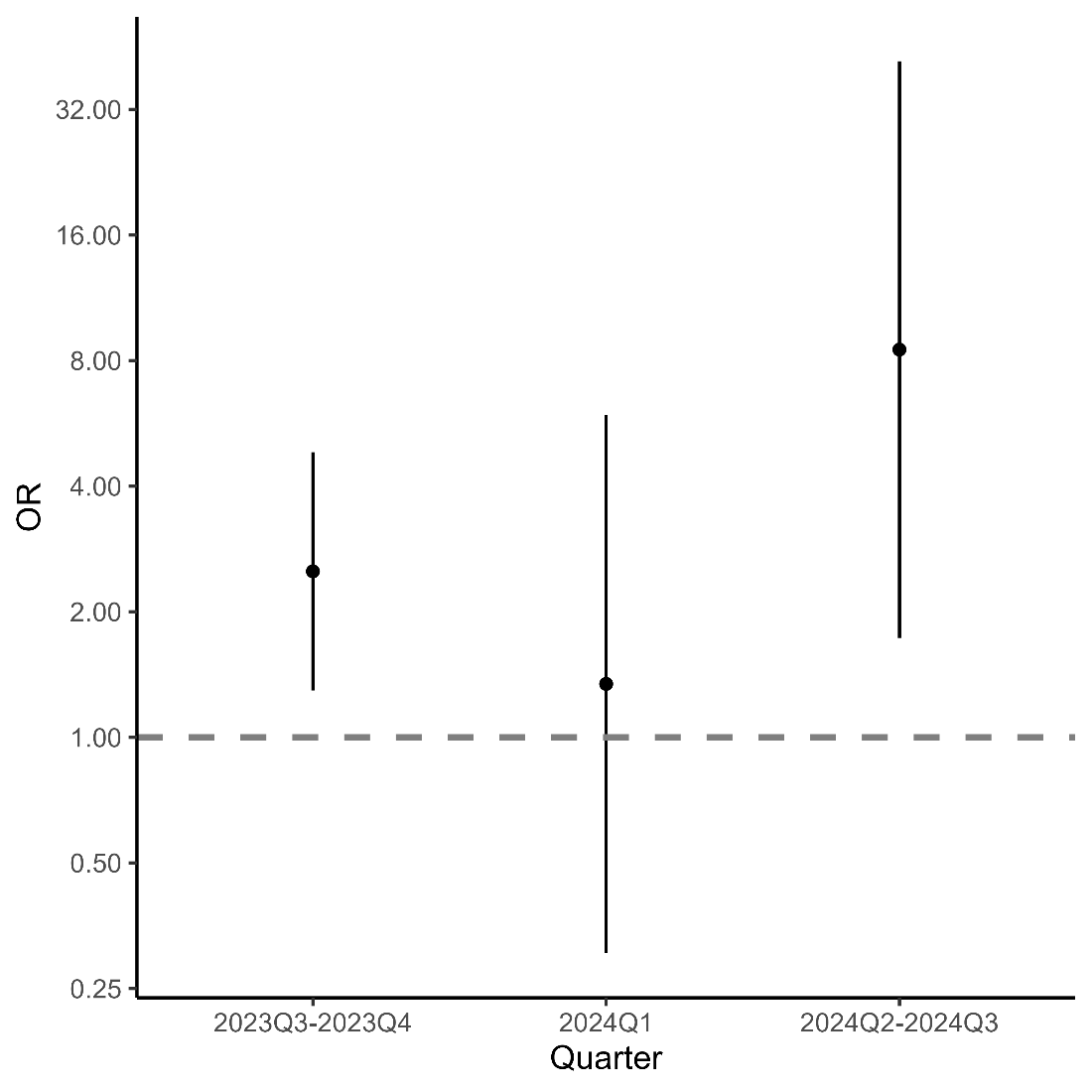

**Supplemental Figure 4.** Relationship between travel to areas of higher incidence (ORs and 95% CI) over time, with only confirmed malaria cases as outcome.

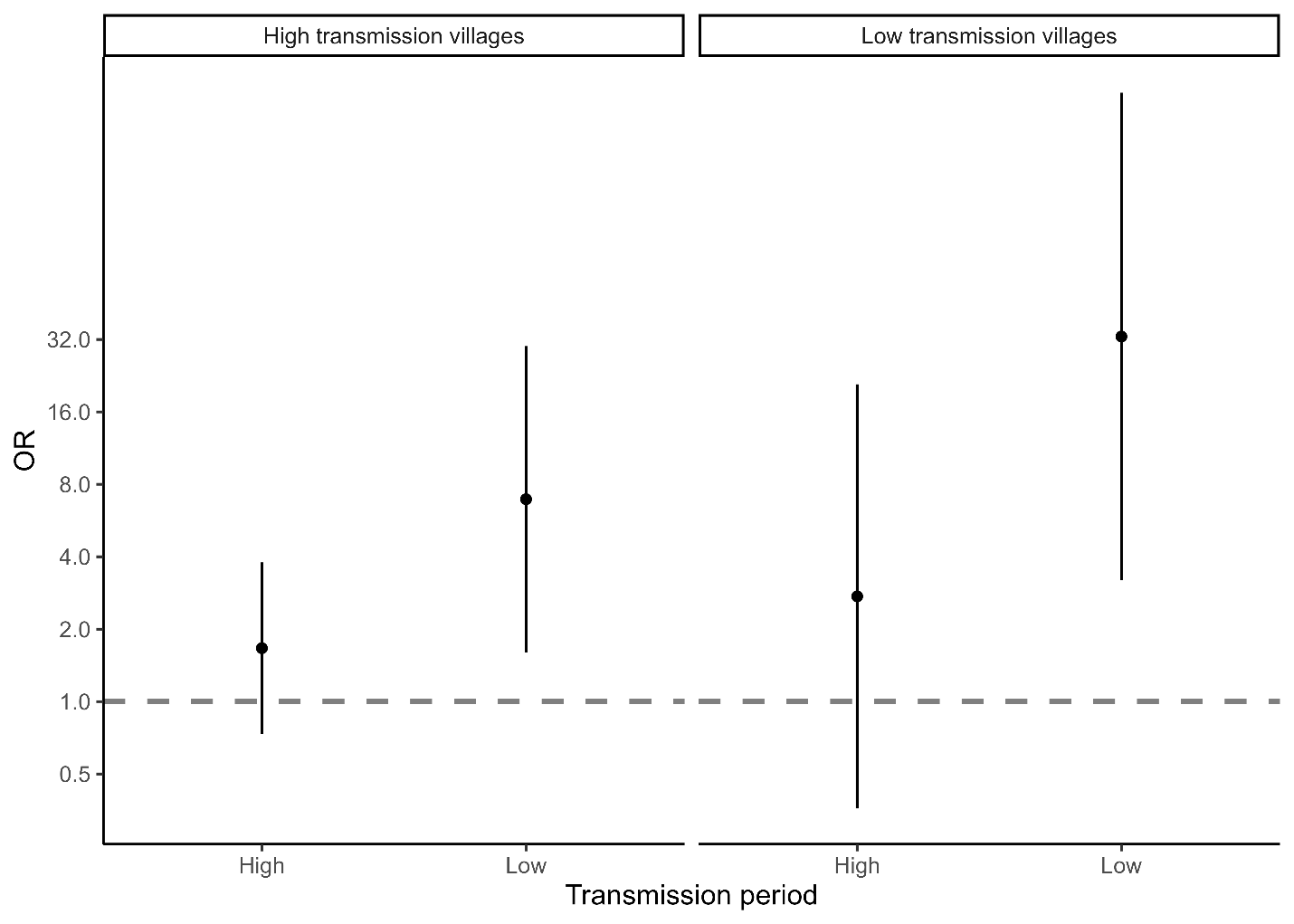

**Supplemental Figure 5.** Relationship between travel to areas of higher incidence (ORs and 95% CI) by time and village-level transmission intensity, with only confirmed malaria cases as outcome.
